# Factors associated with poor glycaemic control among children and adolescents with Type 1 Diabetes Mellitus at a tertiary hospital in Kenya

**DOI:** 10.64898/2026.10.14.26365593

**Authors:** Anjumanara Omar, Diana Marangu-Boore, Fred Bukachi, Fred Were

**Author notes:** Corresponding author, (AO).

## Abstract

Type 1 diabetes mellitus (T1DM) remains a growing global health priority, with disproportionate underdiagnosis, delayed diagnosis, and mortality in sub-Saharan Africa. Despite the introduction of the Life for a Child (LFaC) programme in Kenya, which provides free insulin, glucose-monitoring supplies, and specialist education, no published studies have examined the determinants of glycaemic control among Kenyan children and adolescents with T1DM. This study aimed to determine the proportion of children and adolescents achieving good glycaemic control and to identify the socio-demographic and clinical determinants of glycaemic control at a tertiary hospital in Kenya.

This cross-sectional study enrolled 160 children and adolescents with T1DM attending the paediatric diabetes clinic at Kenyatta National Hospital, Nairobi, between September 2025 and May 2026. Sociodemographic and clinical data were collected through structured interviews and physical examination. Poor glycaemic control was defined as HbA1c >7.5%. Associations between candidate determinants and glycaemic control were assessed using chi-square or Fisher’s exact tests. Variables with p<0.2 on bivariate analysis were entered into a multivariable logistic regression model; Firth’s penalized-likelihood method was used given the small number of participants achieving good control.

Only 9 participants (5.6%) achieved the target HbA1c of ≤7.5% (mean HbA1c 11.02%, SD ±2.29), and only 5 of 133 patients (3.8%) had ≥70% of glucometer readings within the target blood glucose range (3.9–10.0 mmol/L).On bivariate analysis, household income (p=0.026) and self-monitored blood glucose (SMBG) frequency (p=0.018) were significantly associated with glycaemic control. In an exploratory multivariable regression analysis (Firth penalized logistic regression, given the small number of participants with good control), only SMBG frequency ≥4 tests/day remained independently associated with lower odds of poor control (adjusted OR 0.19, 95% CI 0.04–0.84, p=0.028); the effect of household income was attenuated after adjustment.

Glycaemic control remains markedly poor among children and adolescents with T1DM in this Kenyan tertiary hospital setting despite structured programmatic support. Frequency of self-monitoring, rather than household income alone, was the factor most strongly associated with glycaemic control, suggesting that strengthening access to glucose-testing supplies and supporting their consistent use should be prioritised in care, alongside attention to socioeconomic barriers. Longitudinal studies are needed to confirm these associations.

---

Type 1 diabetes mellitus (T1DM) is the most common endocrine disorder of childhood and one of the leading chronic diseases affecting children and adolescents worldwide. It results from autoimmune destruction of pancreatic β-cells, leading to absolute insulin deficiency, with disease onset occurring in genetically susceptible individuals following exposure to environmental triggers [1]. Global estimates indicate that the number of individuals living with T1DM reached approximately 9.2 million in 2024, of whom an estimated 1.8 million are under the age of 20 years; a further two million individuals with T1DM reside in low-income or lower-middle-income countries [2,3]. The increasing incidence, coupled with widening disparities in access to insulin, diabetes technologies, and specialist care, has made T1DM an emerging global health priority, particularly in sub-Saharan Africa where underdiagnosis, delayed diagnosis, and increased mortality remain common [4]. Kenya reflects this burden closely: national estimates indicate that 570 children and adolescents died from T1DM in 2024 alone, with approximately 6,500 children currently living with the condition and 1,380 new diagnoses that year [5]. Local outcome data remain equally sparse: the only published Kenyan study on this population, by Ngwiri et al., carried out across three outpatient clinics in the Nairobi area in 2015, found a median HbA1c of 11.1% and reported that only 28% of children and adolescents achieved reasonable glycaemic control, with age above 12 years significantly associated with poorer control [6].

Insulin therapy is indispensable for survival in T1DM, and frequent blood glucose monitoring is essential to achieving optimal glycaemic targets [7]. It is well established that inadequate glycaemic control substantially increases the risk of both acute and chronic diabetes-related complications [8]. Glycaemic control is conventionally assessed by glycated haemoglobin (HbA1c), which reflects mean plasma glucose over approximately three months, and by Time in Range (TIR), defined as the proportion of time spent within the target glucose range of 3.9–10.0 mmol/L (70–180 mg/dL) [9]. A TIR of 70% or greater is the internationally recommended clinical target [10].

The International Society for Paediatric and Adolescent Diabetes (ISPAD) recommends an HbA1c target of ≤6.5% (48 mmol/mol) for individuals utilising advanced technologies including Continuous Glucose Monitoring (CGM), and ≤7.0% (53 mmol/mol) in all other circumstances. However, ISPAD and the American Diabetes Association (ADA) acknowledge that less stringent targets of ≤7.5% may be appropriate in selected populations, particularly in resource-constrained settings where hypoglycaemia risk cannot be adequately monitored [11,12].

Evidence from the Diabetes Control and Complications Trial (DCCT) and its Epidemiology of Diabetes Interventions and Complications (EDIC) follow-up revealed that intensive glycaemic control initiated during childhood and adolescence confers long-term protection against microvascular and macrovascular complications, even when subsequent glycaemic control converges between treatment groups [8,13].

Although poor glycaemic control among children and adolescents with T1DM has been constantly documented across low- and middle-income countries, the determinants identified have varied considerably between settings. While studies from Ethiopia and Tanzania highlight socioeconomic deprivation and limited access to diabetes technologies, others identify behavioral factors, caregiver engagement, health literacy, insulin availability, and health system organization as the main factors. This heterogeneity suggests that determinants of glycaemic control are highly context-specific and cannot be generalized across health systems without local evidence [14,15,16].

The introduction of the Life for a Child (LFaC) programme has substantially reduced barriers to essential diabetes commodities. However, improved access to insulin and glucose monitoring does not necessarily translate into improved metabolic outcomes [17]. Evidence from several LMICs demonstrates that glycaemic control remains suboptimal despite enhanced access to diabetes supplies, highlighting the importance of behavioural, caregiver, psychosocial, and health-system determinants [18].

Despite increasing access to diabetes care through the LFaC programme, no published studies have examined the determinants of glycaemic control among Kenyan children and adolescents with T1DM. Consequently, it remains unclear which socio-demographic and clinical factors are associated with glycaemic outcomes in this population.

Therefore, this study aimed to determine the proportion of children and adolescents achieving good glycaemic control and to identify the socio-demographic and clinical determinants of glycaemic control at a tertiary hospital in Kenya. Factors were grouped according to the conceptual framework shown in Fig 1

**Fig 1.**
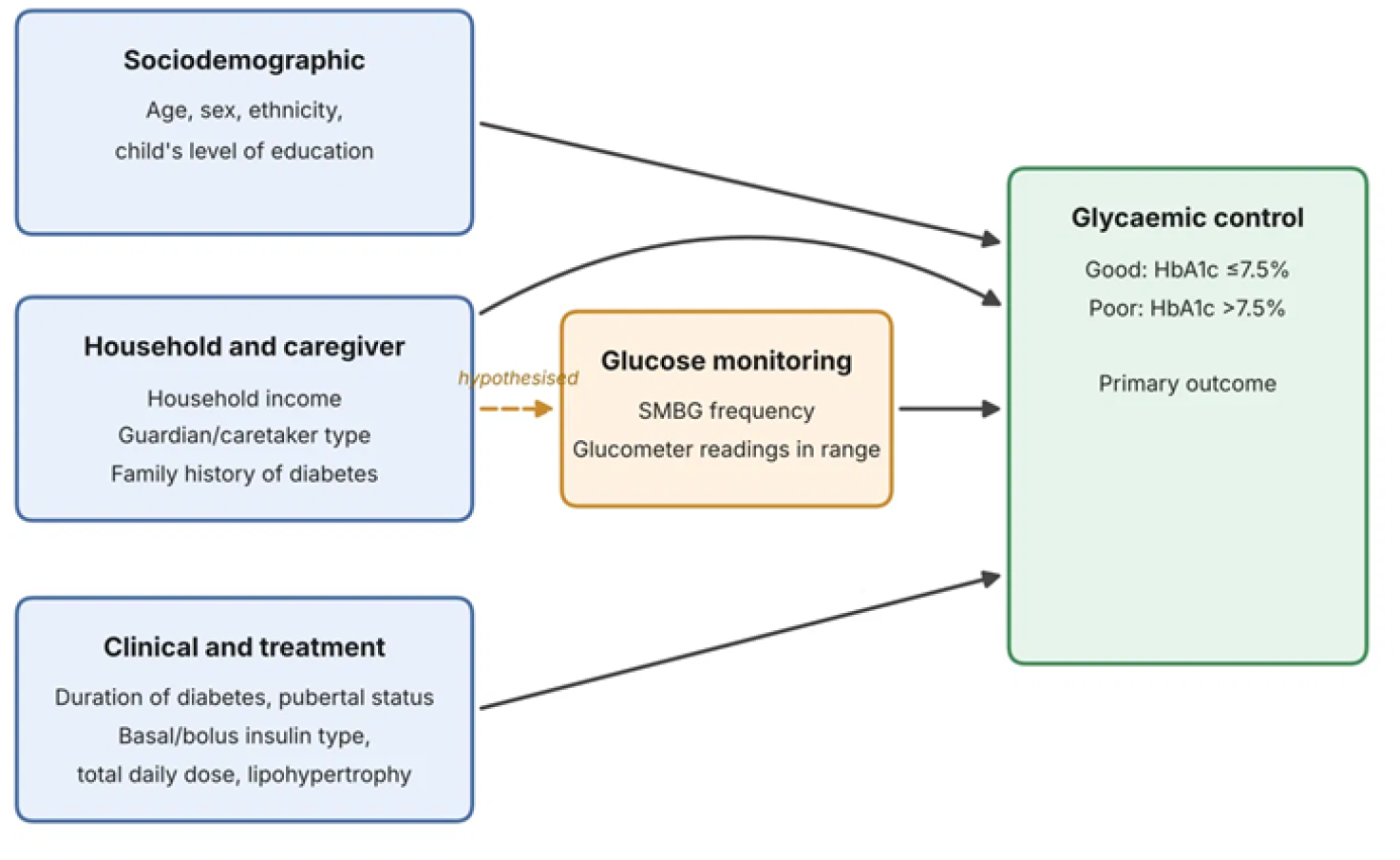
Conceptual framework of factors associated with glycaemic control. Arrows show hypothesised associations; the dashed arrow shows the hypothesised pathway from household socioeconomic status through glucose monitoring.

## METHODS

### Study Setting and Design

This cross-sectional study was conducted at the paediatric diabetes clinic at Kenyatta National Hospital (KNH), Nairobi, which is the largest national referral hospital in Kenya, with an inpatient capacity of 1,800 beds. The clinic serves a catchment population extending across all counties of Kenya and is believed to manage the majority of children and adolescents with T1DM in the country. The clinic is administered by a multidisciplinary team comprising paediatric endocrinologists, endocrinology fellows, paediatric residents, and two certified paediatric diabetes nurse educators.

Participants were recruited consecutively during routine clinic visits between September 2025 and May 2026 and data was collected at the time of each visit using a structured questionnaire.

### Patient Population and Clinical Protocol

All patients with T1DM are initiated on a basal-bolus insulin regimen comprising glargine and regular insulin at enrolment, with doses titrated according to weight, age, and prevailing blood glucose values. Patients and caregivers receive structured diabetes education at each clinic attendance. A glucometer is provided at the point of initiation of care, and patients are reviewed at three-monthly intervals with sustained access to glucose monitoring strips. Glycaemic control is evaluated at each visit using HbA1c and glucometer based target glucose range.

### Eligibility Criteria

All children and adolescents diagnosed with T1DM in accordance with ISPAD diagnostic criteria who had attended at least two clinic evaluations were eligible for inclusion. Signed written informed consent was obtained from the parent or legal guardian of all participants under 18 years of age; verbal assent was additionally obtained from participants aged 7 years and above. Ethical approval was granted by the KNH–University of Nairobi Ethics and Research Committee.

### Conceptual Framework

Factors of glycaemic control were selected from the literature and grouped into four domains: sociodemographic, household and caregiver, clinical and treatment-related, and monitoring-related factors (Fig 1). Household socioeconomic status was hypothesised to influence glycaemic control both directly and through the frequency of glucose monitoring. Because of the cross-sectional design, the framework describes hypothesised associations and not causal pathways.

### Data Collection

Sociodemographic and clinical data were obtained through structured face-to-face interviews and physical examination. Anthropometric measurements were taken using a calibrated digital scale (accurate to 0.1 kg) for weight and a vertical stadiometer (accurate to 0.1 cm) for height; body mass index (BMI) was calculated and plotted on CDC gender-specific growth charts. All participants underwent physical examination for injection-site lipodystrophy and pubertal staging according to Tanner criteria.

### Glycaemic Assessment

Venous blood samples were analysed for HbA1c using the iChroma II point-of-care device, which yields results expressed as a percentage. The device was calibrated against IFCC/NGSP-standardised reference methods. Poor glycaemic control was defined as HbA1c >7.5%, consistent with the less stringent target endorsed by ISPAD and ADA for resource-limited settings [10,12]. Glycaemic control within the target glucose range was assessed from glucometer data in the 133 patients who presented with their device at the clinic visit. The Accu-Chek glucometer (Roche Diabetes Care) was connected to a computer, and stored readings were downloaded and analysed using Accu-Chek Smart Pix software, which reports the proportion of readings within the target range (3.9–10.0 mmol/L). The software reporting period was set to three months, the longest available option, which corresponds to the period reflected by HbA1c. A value of ≥70% of readings in range was defined as desirable, by analogy with the international consensus target for CGM-derived time in range [11]. This measure is referred to as “readings in range” throughout. It should be interpreted as a non-validated proxy, because intermittent fingerstick readings do not replicate the continuous interstitial glucose data on which time-in-range definitions and clinical targets are based. This measure is referred to as “readings in range” throughout, and it should be interpreted as a non-validated proxy: intermittent fingerstick readings do not replicate the continuous interstitial glucose data on which time-in-range definitions and clinical targets are based, and they may over-represent times when testing was prompted by symptoms or by meals [11]

### Sample Size Determination

The sample size was adequate for estimating the prevalence of poor glycaemic control from a comparable African setting [19] but was not necessarily powered to detect associations in multivariable analysis.

Using a 95% confidence interval and a 5% margin of error, with a 10% allowance for non-response, the minimum required sample was 136 participants. A total of 160 patients were enrolled, exceeding this minimum.

Potential sources of bias included selection bias, as only patients attending KNH Paediatric Endocrine clinic and having completed at least two clinic visits were eligible for inclusion, and measurement bias due to the use of glucometer-derived glucose readings rather than TIR from continuous glucose monitoring. Standardized data collection procedures and HbA1c measurements were used to minimise these biases.

### Statistical Analysis

Data were coded, cleaned, and analysed using SPSS version 26.0 (IBM Corporation, Armonk, NY). Categorical variables were summarised as frequencies and percentages; continuous variables as means with standard deviations or medians with interquartile ranges, as appropriate to the distribution. Bivariate analysis was conducted using chi-square or Fisher’s exact tests to examine associations between potential determinants and glycaemic control, with statistical significance set at p < 0.05. Although a multivariable logistic regression model was explored, the small number of patients achieving good glycaemic control (n = 9; 5.6%) precluded stable estimation, as this fell below the recommended minimum of ten events per predictor variable; the multivariable analysis was therefore presented as exploratory and hypothesis-generating only. Analyses were conducted using available-case data; no attribution was performed for missing values.

## RESULTS

### Sociodemographic Characteristics

A total of 160 children and adolescents with T1DM were enrolled. The mean age was 12.0 years (SD ±4.09; median 12.1 years, IQR 9.2–15.1), with 48.1% of participants being female. The Kikuyu ethnic group constituted the majority (57.5%). Caretaker arrangement was predominantly dual-parental (71.9%). Primary school was the most commonly attended level of education (52.5%), followed by secondary school (29.4%). Household income was self-reported by the caregiver and categorized into predefined income brackets. Detailed sociodemographic characteristics are presented in Table 1.

**Table 1.** Sociodemographic characteristics of children and adolescents with type 1 diabetes mellitus (n = 160). KES = Kenyan Shillings; SD = standard deviation; IQR = interquartile range.

| Variable | Category | Frequency (n=160) | Percentage (%) |
| --- | --- | --- | --- |
| **Age (years)**<br>**Mean 12.0 years (SD $\pm 4.09$);**<br>**Median 12.1 years (IQR 9.2–15.1)** | $\leq 5.0$ | 10 | 6.3 |
|  | 5.1–12.0 | 69 | 43.1 |
| | $\geq 12.1$ | 81 | 50.6 |
| Gender | Female | 77 | 48.1 |
|  | Male | 83 | 51.9 |
| Ethnicity | Kikuyu | 92 | 57.5 |
|  | Kamba | 18 | 11.3 |
|  | Somali | 13 | 8.1 |
|  | Luhya | 10 | 6.3 |
|  | Luo | 7 | 4.4 |
|  | Maasai | 6 | 3.8 |
|  | Other | 14 | 8.8 |
| Education level of child | None | 13 | 8.1 |
|  | Nursery | 10 | 6.3 |
|  | Primary | 84 | 52.5 |
|  | Secondary | 47 | 29.4 |
|  | Tertiary | 6 | 3.8 |
| Guardian/caretaker | Both parents | 115 | 71.9 |
|  | Single parent | 40 | 25.0 |
|  | Caregiver (non-parent) | 5 | 3.1 |
| Household income per month (KES) | Below 24,000 | 58 | 36.3 |
|  | 24,000–<100,000 | 88 | 55 |
|  | >100,000 | 14 | 8.8 |
| Family history of diabetes mellitus | Yes | 54 | 33.8 |
|  | No | 106 | 66.3 |

### Clinical and Treatment-Related Characteristics

The mean duration of T1DM was 4.45 years (SD ±3.27). Of the 158 participants whose bolus insulin data was available, regular insulin was more frequently used (66.5%) than rapid-acting analogs (33.5%). As for the basal insulin, glargine was the most common formulation used at 91.3%. More than half of the cohort (55.0%, n = 88) were pubertal at the time of assessment. Among the 160 participants, 151 (94.4%) had poor glycaemic control (HbA1c >7.5%), while only 9 (5.6%) achieved the target HbA1c of ≤7.5%. Glucometer-derived readings in range were assessed in 133 participants who presented with their glucometer. Of these, only 5 (3.8%) had ≥70% of readings within the target blood glucose range (3.9–10.0 mmol/L), while 128 (96.2%) had <70% of readings in range. Self-monitored blood glucose (SMBG) frequency data was available for 136 participants, of whom 90 (66.2%) tested fewer than 4 times per day, and 46 (33.8%) tested 4 or more times per day. The mean HbA1c was 11.02% (SD ± 2.29), with a median of 10.7% (IQR 9.60–12.67). Full clinical and treatment-related characteristics are shown in Table 2.

**Table 2.** Clinical and treatment-related characteristics of children and adolescents with type 1 diabetes mellitus (n = 160, unless otherwise stated). HbA1c = glycated haemoglobin; RIR = Readings in range: percentage of glucometer readings within 3.9–10.0 mmol/L over the preceding three months, from Accu-Chek Smart Pix; SMBG = self-monitored blood glucose; TDD = total daily dose.

| Variable | Category | Frequency (n) | Percentage (%) |
| --- | --- | --- | --- |
| HbA1c (%)<br>Mean 11.02% (SD ±2.29); Median 10.7% (IQR 9.60–12.67) | >7.5% | 151 | 94.4 |
|  | ≤7.5% | 9 | 5.6 |
| Basal insulin | Glargine | 146 | 91.3 |
|  | Detemir | 13 | 8.1 |
|  | Premix 70/30 | 1 | 0.6 |
| Bolus insulin (n = 158) | Regular insulin | 105 | 66.5 |
|  | Rapid-acting (Aspart/Lispro) | 53 | 33.5 |
| RIR (n = 133) | ≥70% | 5 | 3.8 |
|  | <70% | 128 | 96.2 |
| SMBG frequency (n = 136) | <4 tests per day | 90 | 66.2 |
|  | ≥4 tests per day | 46 | 33.8 |
| Lipohypertrophy | Absent | 121 | 75.6 |
|  | Present | 39 | 24.4 |
| Pubertal status | Prepubertal | 72 | 45.0 |
|  | Pubertal | 88 | 55.0 |
| TDD (units/kg/day) | <1.00 | 141 | 88.1 |
|  | ≥1.00 | 19 | 11.9 |
| Duration of diabetes | ≤5 years | 100 | 62.5 |
| Mean 4.5 years (SD ±3.3) | >5 years | 60 | 37.5 |

### Sociodemographic factors associated with Glycaemic Control

In bivariate analysis, household income was the only sociodemographic variable significantly associated with poor glycaemic control (p=0.026). Among patients with HbA1c ≤7.5%, 33.3% had household incomes exceeding KES 100,000 per month, compared with only 7.3% in the poor glycaemic control group, suggesting a relationship between higher socioeconomic status and better glycaemic outcomes. No statistically significant associations were identified between glycaemic control and age, sex, family history of diabetes mellitus, or guardian/caretaker type, although a non-significant trend toward better control was observed among children with a positive family history of diabetes (p=0.167) as shown in Table 3.

**Table 3.**
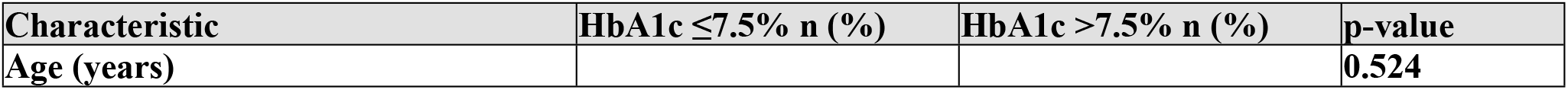
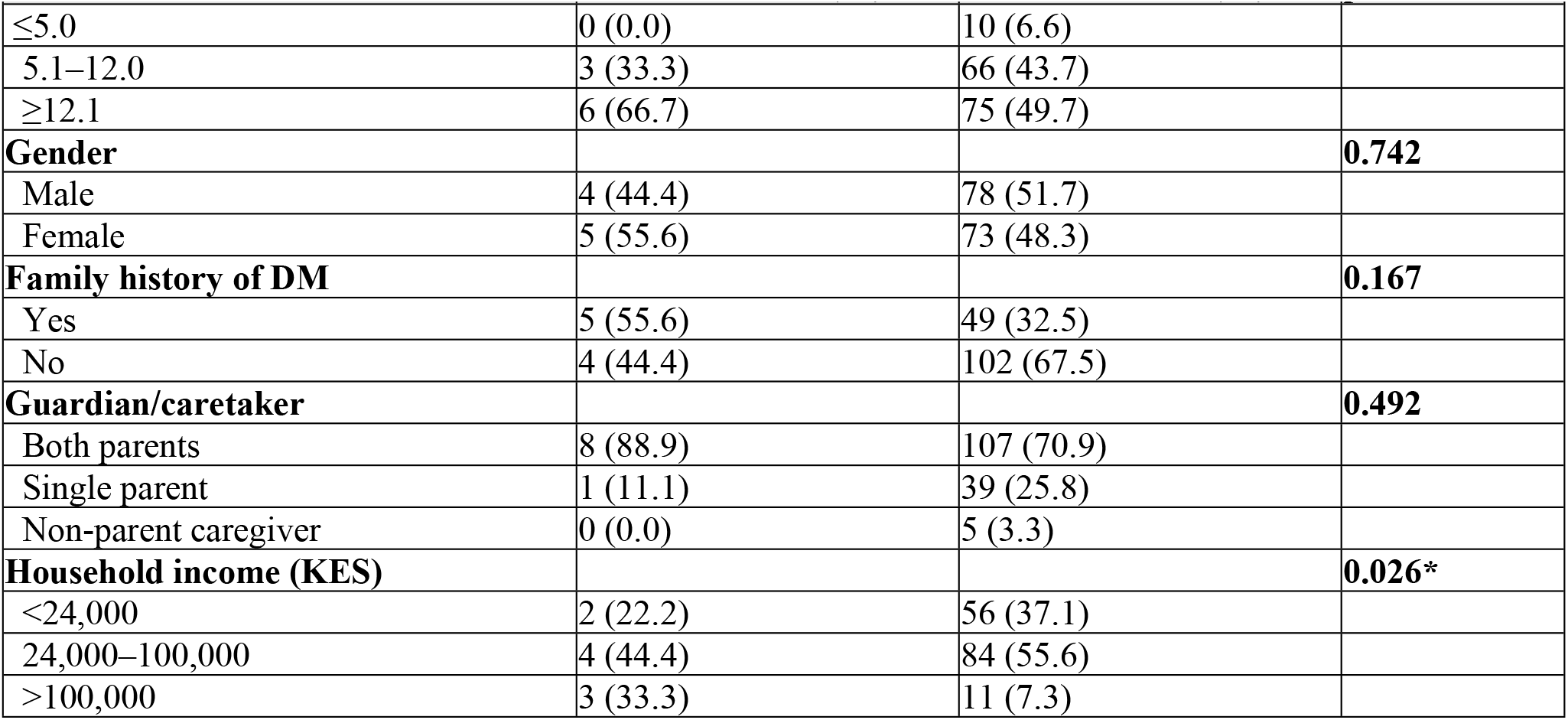
Bivariate analysis of sociodemographic factors associated with glycaemic control in children and adolescents with T1DM (n = 160). * Statistically significant at p < 0.05. KES = Kenyan Shillings. Fisher’s exact test applied where expected cell counts were <5.

### Clinical and Treatment-Related Factors Associated with Glycaemic Control

None of the clinical or treatment-related characteristics examined, including bolus and basal insulin type, total daily insulin dose, lipohypertrophy, pubertal status, glucometer-derived readings in range, or duration of diabetes, were significantly associated with glycaemic control in bivariate analysis, as presented in Table 4. However, SMBG was significantly associated with poor glycaemic control (p=0.018): 75.0% of participants with good control tested ≥4 times per day, compared with only 31.3% of those with poor control.

**Table 4.**
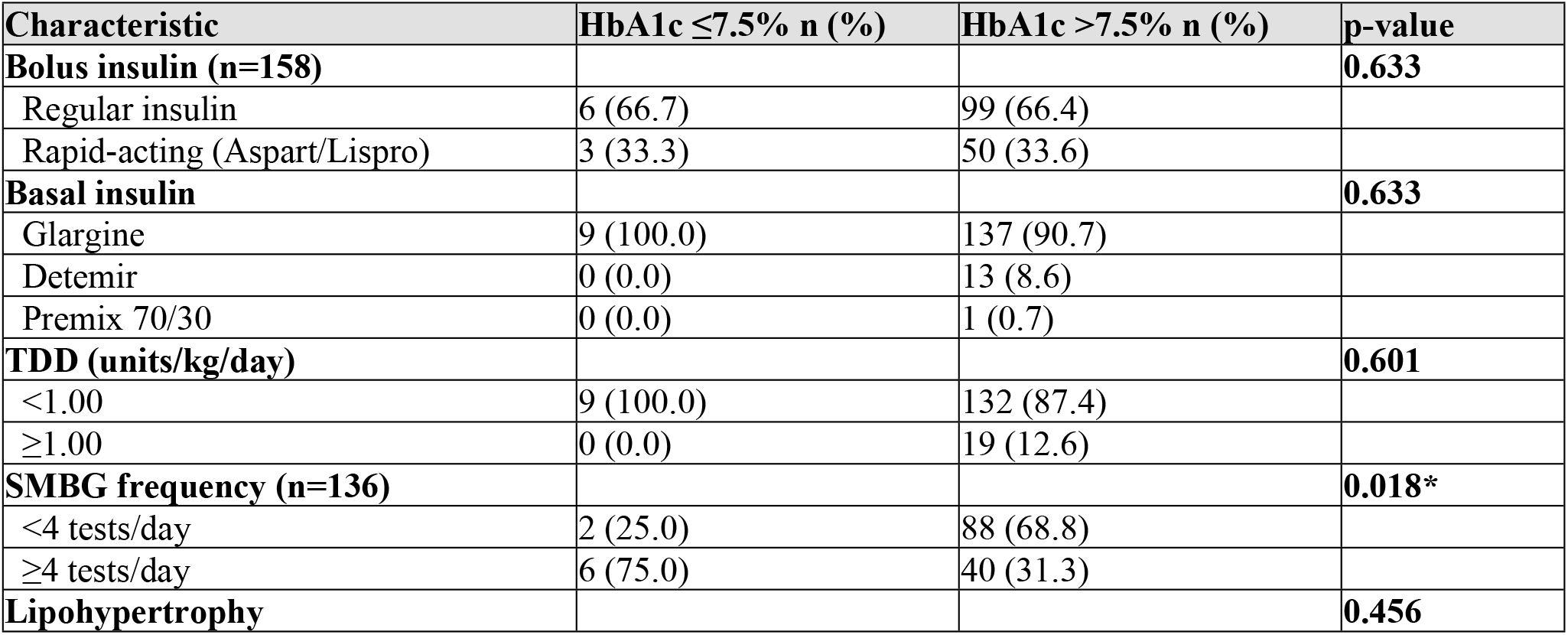
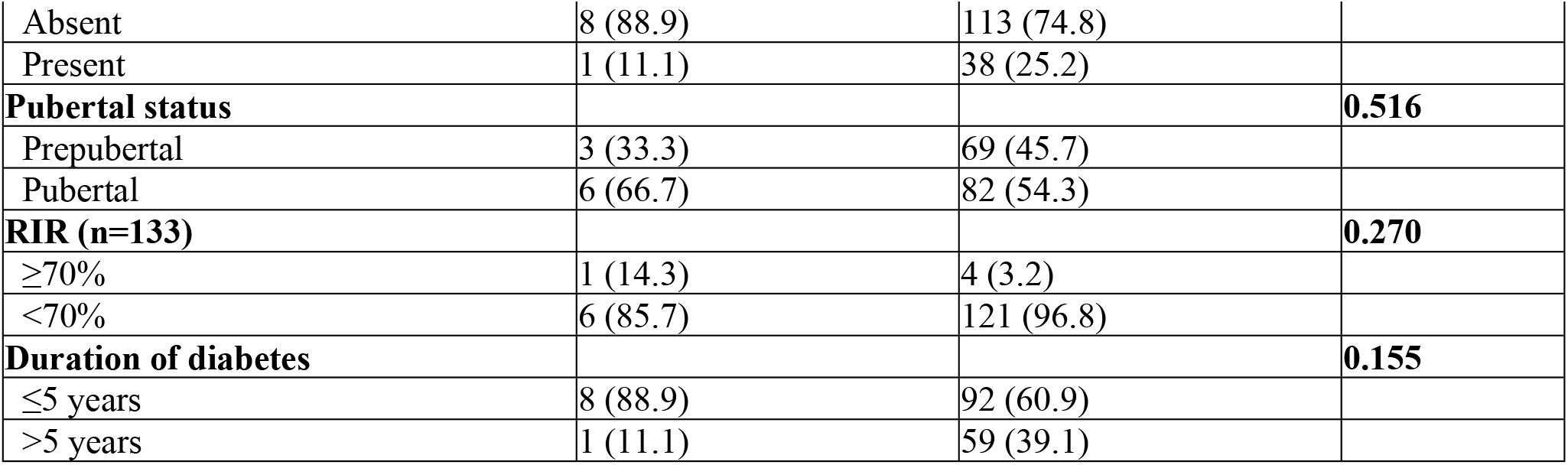
Bivariate analysis of clinical and treatment-related factors associated with glycaemic control (n = 160, unless otherwise stated). *Statistically significant at p<0.05. Fisher’s exact test applied where expected cell counts were <5. SMBG = self-monitored blood glucose; RIR = Readings in range: percentage of glucometer readings within 3.9–10.0 mmol/L over the preceding three months, from Accu-Chek Smart Pix; TDD = total daily dose.

| Characteristic | HbA1c ≤7.5% n (%) | HbA1c >7.5% n (%) | p-value |
| --- | --- | --- | --- |
| **Bolus insulin (n=158)** |  |  | **0.633** |
| Regular insulin | 6 (66.7) | 99 (66.4) |  |
| Rapid-acting (Aspart/Lispro) | 3 (33.3) | 50 (33.6) |  |
| **Basal insulin** |  |  | **0.633** |
| Glargine | 9 (100.0) | 137 (90.7) |  |
| Detemir | 0 (0.0) | 13 (8.6) |  |
| Premix 70/30 | 0 (0.0) | 1 (0.7) |  |
| **TDD (units/kg/day)** |  |  | **0.601** |
| <1.00 | 9 (100.0) | 132 (87.4) |  |
| ≥1.00 | 0 (0.0) | 19 (12.6) |  |
| **SMBG frequency (n=136)** |  |  | **0.018\*** |
| <4 tests/day | 2 (25.0) | 88 (68.8) |  |
| ≥4 tests/day | 6 (75.0) | 40 (31.3) |  |
| **Lipohypertrophy** |  |  | **0.456** |
| Absent | 8 (88.9) | 113 (74.8) |  |
| Present | 1 (11.1) | 38 (25.2) |  |
| **Pubertal status** |  |  | **0.516** |
| Prepubertal | 3 (33.3) | 69 (45.7) |  |
| Pubertal | 6 (66.7) | 82 (54.3) |  |
| **RIR (n=133)** |  |  | **0.270** |
| ≥70% | 1 (14.3) | 4 (3.2) |  |
| <70% | 6 (85.7) | 121 (96.8) |  |
| **Duration of diabetes** |  |  | **0.155** |
| ≤5 years | 8 (88.9) | 92 (60.9) |  |
| >5 years | 1 (11.1) | 59 (39.1) |  |

After adjustment using Firth penalized-likelihood logistic regression (n=136), SMBG frequency remained the only variable independently associated with glycaemic control: participants who tested ≥4 times per day had significantly lower odds of poor glycaemic control compared with those testing <4 times per day (adjusted OR 0.19, 95% CI 0.04–0.84, p=0.028). Household income, which was significant on bivariate analysis, was no longer independently associated with glycaemic control after adjustment (adjusted OR 1.59, 95% CI 0.30–8.46, p=0.586 for income 24,000–100,000 KES; adjusted

OR 0.25, 95% CI 0.04–1.75, p=0.165 for income >100,000 KES, both relative to income <24,000 KES), suggesting that its bivariate association may partly operate through testing frequency, although the small number of participants with good control limits the precision of these estimates. Family history of diabetes was not independently associated with glycaemic control (adjusted OR 0.50, 95% CI 0.12– 2.15, p=0.355). Results are presented in Table 5

**Table 5:** Univariable and multivariable analysis of determinants of glycaemic control Statistically significant at p<0.05. Variables with crude p<0.2 (household income, SMBG frequency, family history of diabetes) were entered into the multivariable model (n=136). Firth’s penalized-likelihood logistic regression was used given the small number of participants with good glycaemic control (n=9), which precluded stable estimation using conventional maximum-likelihood logistic regression. Dashes indicate variables not entered into the multivariable model. KES = Kenyan Shillings; SMBG = self-monitored blood glucose; RIR = Readings in range: percentage of glucometer readings within 3.9–10.0 mmol/L over the preceding three months, from Accu-Chek Smart Pix; TDD = total daily dose

| Determinant | Crude OR | 95% CI | p-value | Adjusted OR | 95% CI | p-value |
| --- | --- | --- | --- | --- | --- | --- |
| Age (years) | 0.96 | 0.82–1.11 | 0.568 | — | — | — |
| Duration of diabetes (years) | 1.17 | 0.90–1.51 | 0.236 | — | — | — |
| Sex, male (ref: female) | 1.32 | 0.36–4.83 | 0.671 | — | — | — |
| Income 24,000–100,000 KES (ref: <24,000) | 0.83 | 0.17–4.09 | 0.820 | 1.59 | 0.30–8.46 | 0.586 |
| Income >100,000 KES (ref: <24,000) | 0.15 | 0.03–0.86 | 0.034* | 0.25 | 0.04–1.75 | 0.165 |
| Guardian, single parent (ref: both parents) | 2.08 | 0.35–12.52 | 0.423 | — | — | — |
| Guardian, non-parent caregiver (ref: both parents) | 0.87 | 0.03–22.39 | 0.933 | — | — | — |
| Family history of DM (ref: no) | 0.40 | 0.11–1.45 | 0.162 | 0.50 | 0.12–2.15 | 0.355 |
| Pubertal (ref: prepubertal) | 0.64 | 0.17–2.46 | 0.515 | — | — | — |
| Lipohypertrophy present (ref: absent) | 1.92 | 0.32–11.56 | 0.475 | — | — | — |
| TDD ≥1.00 units/kg/day (ref: <1.00) | 2.80 | 0.15–53.69 | 0.495 | — | — | — |
| SMBG ≥4 tests/day (ref: <4) | 0.18 | 0.04–0.80 | 0.025* | 0.19 | 0.04–0.84 | 0.028* |
| RIR ≥70% (ref: <70%) | 0.19 | 0.02–1.60 | 0.125 | — | — | — |

## DISCUSSION

This cross-sectional study of 160 children and adolescents with T1DM attending the KNH paediatric diabetes clinic demonstrates that glycaemic control remains markedly poor despite the availability of a structured support programme that includes free insulin, glucose-monitoring supplies, and specialist diabetes education at every visit. Only 9 participants (5.6%) achieved the study-defined HbA1c target of ≤7.5%, and a mere 5 of 133 patients assessed (3.8%) had ≥70% of glucometer readings within the target blood glucose range. In bivariate analysis, low household income (p = 0.026) and infrequent self-monitoring of blood glucose (p = 0.018) were significantly associated with poor glycaemic control, and in exploratory multivariable analysis, only SMBG frequency remained independently associated with poor control.

The prevalence of poor glycaemic control observed in this cohort (94.4%) with HbA1c >7.5% is consistent with findings from comparable sub-Saharan African settings. Ogugua et al. documented poor glycaemic control in 78.9% of children with T1DM in South Eastern Nigeria [19].Similarly, Ali and Alhassan reported that only 6.2% of children with T1DM in Sudan achieved adequate glycaemic control [20], and Muchunguzi et al. reported suboptimal control in 66.4% of patients in Tanzania [21]. The median HbA1c of 10.7% (mean 11.02%) in the present study is similar to the median of 11.1% reported by Ngwiri et al. across three Nairobi facilities, including KNH [6]. This suggests that glycaemic outcomes may have changed little over the intervening decade, despite the establishment of specialist clinic infrastructure and the introduction of the LFaC supply programme, although differences in study populations and sampling limit direct comparison.

The clinical consequences of sustained poor glycaemic control are well characterized. McCarter et al. demonstrated that elevated HbA1c is associated with a threefold increase in the risk of retinopathy and a sixfold increase in the risk of nephropathy in T1DM [22]. The DCCT/EDIC cohort established definitively that intensive glycaemic control significantly reduces the incidence and progression of microvascular complications, particularly retinopathy, nephropathy, and neuropathy, with benefits that persist for decades following the period of intensive intervention [8,23]. In a cohort where the mean HbA1c exceeds 11%, the cumulative microvascular burden accruing over childhood and adolescence is likely to be substantial. The broader systemic context characterized by unreliable insulin supply chains, limited specialist access, and high rates of DKA at presentation compounds this risk [24,25].

The significant association between low socioeconomic status and poor glycaemic control (p = 0.026) is consistent with a robust body of international evidence. Bijlsma-Rutte et al. demonstrated, in a systematic review and meta-analysis, a well-established inverse relationship between socioeconomic status and HbA1c in diabetes [26]. Sebastian-Valles et al. similarly documented worse glycaemic outcomes among individuals with T1DM from lower socioeconomic strata, even when flash glucose systems were in use [27].

Self-monitored blood glucose (SMBG) frequency was similarly associated with glycaemic control in this cohort, on both bivariate and exploratory multivariable analyses. Ziegler et al., in a large registry-based study, found that SMBG frequency correlated with both HbA1c and the rate of acute complications in children and adolescents with T1DM [28]. A managed-care paediatric practice similarly reported a highly significant decrease in HbA1c as SMBG frequency increased, with each additional daily measurement associated with an average 0.19% reduction in HbA1c [29]. These findings, together with the socioeconomic gradient observed in this study, support the interpretation that the income effect on glycaemic control may operate substantially through its influence on testing capacity. One possible explanation would be that higher-income households may have greater capacity to sustain glucose-testing supplies

In the Kenyan context, low socioeconomic status confers multiple compounding disadvantages: the cost of transport from rural and peri-urban areas to KNH may impede clinic attendance; food insecurity and irregular meal patterns may precipitate missed or reduced insulin doses secondary to hypoglycaemia avoidance; and limited access to reliable electricity in rural households poses a significant challenge to safe insulin storage and preservation. One plausible contributing factor is that when glucose testing strips supplied through the programme are exhausted before the patient’s next scheduled appointment, families may need to purchase replacement strips out-of-pocket, an expense that poorer households may be less able to sustain. This could offer one possible explanation for the observed association between lower income and reduced SMBG frequency in this cohort. These structural determinants operate independently of the availability of insulin and monitoring supplies, explaining why the LFAC programme alone is not sufficient to bridge the glycaemic gap. In support of targeted intervention, Glazier et al., in a systematic review of interventions to improve diabetes care in socially disadvantaged populations, found that intensive, case-based management strategies reduced HbA1c by 1.88% in low-socioeconomic-status groups compared with 1.13% under standard care [30]. These findings strongly endorse the development of community-based, socioeconomically tailored interventions in this population.

Beyond addressing the cost barrier to conventional SMBG, continuous glucose monitoring (CGM) technology offers additional advantages that may be particularly valuable in this population. Unlike intermittent fingerstick testing, CGM provides near-continuous glycaemic data, capturing overnight and postprandial blood sugars that are easily missed with a handful of daily checks, and directly generates Time in Range a metric increasingly favoured over HbA1c alone for its ability to reflect glycaemic variability and hypoglycaemia exposure [31,32]. Given that only 3.8% of participants in this cohort achieved the target blood glucose range despite glucometer availability, broader access to CGM through the LFaC programme could improve both the glycaemic monitoring and clinical decision-making in this setting, independent of household income.

No significant association was identified between glycaemic control and age, sex, ethnicity, educational status, guardian type, or family history of diabetes. These null findings are consistent with previous reports from comparable African settings [33]. The choice of basal or bolus insulin type was likewise not associated with glycaemic outcomes, a finding consistent with the evidence that the clinical benefits of insulin analogues over human insulin are modest in practice and may be attenuated in settings where adherence, monitoring, and titration are constrained by systemic barriers [34,35]. Lipohypertrophy was present in 24.4% of participants but was not significantly associated with HbA1c, consistent with a paediatric Egyptian cohort in which mean HbA1c did not differ by lipohypertrophy grade [36], although lipodystrophy prevalence and its associated factors have also been described in other African paediatric T1DM populations, where insulin injection duration and site-rotation practices, rather than glycaemic control were the primary correlates [37]. Similarly, pubertal status, duration of diabetes, and total daily insulin dose did not reach statistical significance, which is consistent with prior reports in comparable cohorts [16,20].

Despite the widespread implementation of structured support programmes in low- and middle-income countries, optimal glycaemic control remains elusive for the majority of children and adolescents with T1DM, with fewer than 10% achieving target HbA1c levels across most reported cohorts [38,39]. The gap between the goals and outcomes of structured diabetes care is driven by the interplay of systemic barriers, including inadequate healthcare infrastructure, fragmented care pathways, and reliance on supply-focused rather than holistically integrated models and patient-level socioeconomic determinants. Addressing these barriers will require a systems-level response that extends beyond the provision of insulin and monitoring consumables [24,25].

## Limitations

This study has several limitations. Its cross-sectional, single-centre design precludes causal inference and may limit generalisability. Only 9 participants (5.6%) achieved good glycaemic control, so the multivariable analysis was exploratory, with wide confidence intervals. Readings in range were derived from intermittent glucometer data, available for 133 participants, and are a non-validated proxy for CGM-derived time in range. Household income was ascertained by caregiver interview, was unverified and was recorded in broad categories. Data were missing for SMBG (24 participants) and income (8 participants). Finally, adherence, diet, psychosocial and caregiver factors, and the cost of extra testing strips were not measured and may confound the findings.

## Conclusion

Glycaemic control remains markedly poor among children and adolescents with T1DM in this Kenyan tertiary hospital setting, with fewer than 6% achieving target HbA1c despite structured programmatic support, indicating that supply provision alone is insufficient. Higher frequency of self-monitored blood glucose was independently associated with lower odds of poor glycaemic control. Larger, longitudinal studies are needed to confirm these findings and guide future intervention design

## Author Contribution

All authors contributed to the conception and design of the study. Data collection, analysis, and interpretation were performed by the authors. All authors participated in drafting and revising the manuscript and approved the final version.

### Institutional Review Board Statement

This study represents a secondary analysis of data collected as part of a larger cross-sectional study “**Early Indicators Of Cardiovascular Disease In African Children With T1DM”** that received approval by the UoN-KNH Ethics Committee (Approval No. P8/01/2025; approval date: 14/07/2025). No additional participant recruitment or data collection was taken.

### Informed Consent Statement

Written informed consent was obtained from all the legal guardians and assent from all the participants above 7 years prior to participation in the study

## Acknowledgments

The authors would like to thank all the study participants and their families for their participation, as well as the healthcare staff who supported in data collection

## Data Availability Statement

The data underlying the findings of this study contain potentially identifying and sensitive information relating to a paediatric patient population and cannot be made publicly available due to ethical and privacy restrictions imposed by the UoN-KNH Ethics and Research Committee. De-identified data may be made available to qualified researchers upon reasonable request to the corresponding author and with approval from the UoN-KNH Ethics and Research Committee.

### Declaration of generative AI and AI-assisted technologies in the manuscript preparation process

During the preparation of this work, the author used Claude (Anthropic) to assist with statistical analysis, data table generation, and drafting/editing of manuscript text. The authors reviewed and edited the output as needed and take full responsibility for the content of the published article.

